# An infection prediction model developed from inpatient data can predict out-of-hospital COVID-19 infections from wearable data when controlled for dataset shift

**DOI:** 10.1101/2024.12.31.24319816

**Authors:** Ting Feng, Sara Mariani, Bryan Conroy, Robert Damiano, Ikaro Silva, Dennis Swearingen, Daniel C. McFarlane

## Abstract

The COVID-19 pandemic highlighted the importance of early detection of illness and the need for health monitoring solutions outside of the hospital setting. We have previously demonstrated a real-time system to identify COVID-19 infection before diagnostic testing ^1^, that was powered by commercial-off-the-shelf wearables and machine learning models trained with wearable physiological data from COVID-19 cases outside of hospitals. However, these types of solutions were not readily available at the onset nor during the early outbreak of a new infectious disease when preventing infection transmission was critical, due to a lack of pathogen-specific illness data to train the machine learning models. This study investigated whether a pretrained clinical decision support algorithm for predicting hospital-acquired infection (predating COVID-19) could be readily adapted to detect early signs of COVID-19 infection from wearable physiological signals collected in an unconstrained out-of-hospital setting. A baseline comparison where the pretrained model was applied directly to the wearable physiological data resulted a performance of AUROC = 0.52 in predicting COVID-19 infection. After controlling for contextual effects and applying an unsupervised dataset shift transformation derived from a small set of wearable data from healthy individuals, we found that the model performance improved, achieving an AUROC of 0.74, and it detected COVID-19 infection on average 2 days prior to diagnostic testing. Our results suggest that it is possible to deploy a wearable physiological monitoring system with an infection prediction model pretrained from inpatient data, to readily detect out-of-hospital illness at the emergence of a new infectious disease outbreak.

## INTRODUCTION

The COVID-19 pandemic highlighted the importance of early disease detection and isolation in order to prevent the spread of infection ^2–4^. It is desirable, therefore, to have an effective system to continuously monitor an individual’s health state. Health monitoring systems consisting of wearable devices and artificial intelligence (AI) tools are portable, minimally invasive, and were shown to be able to detect COVID-19 infections ^1,5–13^. For example, we developed a real-time infection prediction system using commercial-off-the-shelf (COTS) wearable devices and AI, which was capable of identifying COVID-19 infection on average 2.3 days before diagnostic testing with an Area Under the Receiver Operating Characteristic Curve (AUROC) of 0.82 ^1^. Two other studies reported comparable performance from wearable physiological monitoring with AUROC=0.80 ^5^ and AUROC=0.77 ^14^, respectively.

These health monitoring systems are typically powered by machine learning (ML) models ^1,5–7,14^ or statistical methods ^10^ that are sensitive to physiological changes caused by COVID-19 infection. The models gain intelligence through supervised learning on physiological data collected from the target populations of COVID-19 infection cases. However, training these models require data from a significant number of COVID-19 positive cases, which is challenging because infection data collection is time consuming and costly. Additional challenges of data collection include user compliance, physiologic context effects (such as traveling, intense exercises, etc.), and uncertainties in the timing of infection onset. These models cannot therefore be easily developed when they are most needed, such as at the beginning of novel infection outbreaks like the COVID-19 pandemic.

To this end, we propose that clinical decision support algorithms developed from data collected in hospitals can be utilized to significantly accelerate or provide a minimum viable starting point for wearable systems to monitor for infections in unconstrained, real-world environments. We previously developed a machine learning model that can identify hospital-acquired infection (HAI) patients up to 48 hours before clinical suspicion of infection. The model used physiological measurements from hospital grade devices and demographic information collected in the hospital ^15^. Here, we hypothesized that such infection prediction algorithms trained from hospital dataset (referred to in this article as the “hospital model”) can predict COVID-19 infection from the same set of physiological measurements collected through wearables outside of hospitals, provided that dataset shift ^16^ – the changes in the joint distribution of the physiological features and the infectious disease labels between hospital and wearable datasets – are properly addressed. More specifically, if we define our physiological input features as X and our infectious disease labels as Y, we can typically have three types of dataset shifts:

1. Covariate shift: P(X) changes but P(Y|X) and P(Y) remain the same.
2. Label shift: P(Y) changes but P(Y|X) and P(X) remain the same.
3. Concept drift: P(Y|X) changes but P(X) and P(Y) remain the same.

where P(X), P(Y) and P(Y|X) are the probability distribution of X, probability distribution of Y, and the conditional probability distribution of Y given X, respectively.

In this study, we first performed retrospective analyses to identify sources of dataset shift between hospital dataset and wearable dataset, and then described two correction techniques – removing contextual confounders and applying a monotonic feature transformation – to reduce the differences in data distribution between the two datasets. We found that our infection prediction model trained from the hospital dataset performed best after applying both correction techniques, with an AUROC of 0.74, and detection of COVID-19 infection on average 2 days prior to diagnostic testing. Only a small sample of wearable data from healthy subjects (2 weeks of data from 25 healthy subjects) was required for the feature transformation. Our results suggest that a minimum viable wearable physiological monitoring system that detects early signs of COVID-19 infection can be developed and deployed without the need for data from COVID-19 cases.

## METHODS

### Description of datasets

The two hospital datasets - MIMIC-III (Medical Information Mart for Intensive Care III) ^17^ and Banner Health data - used to train the infection prediction model were described previously ^15^. The two datasets were combined in this study to create a single hospital dataset to train the infection prediction model. Both MIMIC-III and Banner Health data comprise de-identified health-related data from patients during their hospital stay. The MIMIC-III data we used was from patients who stayed in critical care units of the Beth Israel Deaconess Medical Center (Boston, MA) between 2001 and 2012. Each patient encounter included in this study was from the MIMIC-III Waveform Database Matched Subset ^18^. The Banner Health data was from patients who stayed in critical care units or low-acuity settings such as general wards in Banner Health hospitals (Phoenix, AZ). The patient cohort included in this study was collected between 2016 and 2017, where waveform records were available for a subset of the patient encounters.

The wearable dataset used to test hospital model’s ability to detect COVID-19 infection was collected in the framework of a study described previously ^1^ and an extension of the study which focused on algorithm improvement and augmentation. This dataset comprises de-identified COTS wearable physiological and activity data from Garmin watch and Oura ring devices, collected from active military personnel recruited from multiple US Department of Defense (DoD) sites between June 2020 and May 2022. This dataset also included symptoms and diagnostic tests information from self-reported daily survey questionnaires.

### Ethical approval

The MIMIC-III project was approved by the Institutional Review Boards of Beth Israel Deaconess Medical Center and the Massachusetts Institute of Technology (Cambridge, MA). The use of Banner Health data was a part of a retrospective deterioration detection study approved by the Institutional Review Board of Banner Health and by the Philips Internal Committee for Biomedical Experiments. For both hospital datasets, requirements for individual subject consent were waived because the project did not impact clinical care, was no greater than minimal risk, and all protected health information was removed from the limited dataset used in this study.

The collection and use of the wearable dataset was approved by the Institutional Review Boards of the US Department of Defense. Informed consent was obtained from all participants.

### Cohort selection

Patient encounters used in this study to train the hospital-acquired infection prediction model were selected using the same methodology described previously ^15^, namely a set of MIMIC-III and Banner Health patient encounters that had high-sampling frequency waveform recordings around the time of clinical suspicion of hospital-acquired infection. These data were acquired prior to the COVID-19 outbreak therefore did not include instances of COVID-19 infections. We focused on patient encounters with waveform recordings, because we wanted to match the temporal resolution of the vital sign measurements from which the hospital model was trained, with the temporal resolution of the vital sign measurements in the wearable dataset to which the hospital model would be applied. The infection patients, as described previously, were those who had confirmed infection diagnoses and whose timing of clinical suspicion of infection could be localized by a microbiology culture test order. Note that we used as our reference the time when the microbiology culture test was ordered, not the time when the test result was returned. These infection patients were further screened into a hospital-acquired infection cohort if the earliest timing of the microbiology culture test order occurred at least 48 hours after hospital admission.

Subjects used to validate the performance of the hospital model in predicting COVID-19 infection were extracted from the wearable dataset, as described previously ^1^. Specifically, COVID-19 positive subjects were those who reported positive test results and symptoms, and COVID-19 negative subjects were those who reported at least 1 symptom-free negative test result, but no positive results. Condition for inclusion in both classes was the presence of data from a Garmin watch and an Oura ring simultaneously, and that at least 10 nights of physiologic data were collected during sleep within the 21-day period prior to their COVID-19 test (subjects were excluded post-hoc if they did not meet these criteria).

### Feature extraction

The trained hospital model in this study used features derived from a subset of the demographics and vital sign measurements described previously ^15^. We chose this subset because the same set of demographics and vital sign measurements were available and reliable in the wearable dataset. Specifically, the feature vector for training was composed of demographics (age, sex) and four statistic features - average, minimum, maximum and the standard deviation – of core body temperature, respiratory rate, heart rate, and RMSSD (Root Mean Square of Successive Differences between normal heartbeats – a standard measure of heart rate variability), collected in a 24-hour observation window prior to the observation time of 1-hour before clinical suspicion of infection. This resulted in a total set of 18 features in the feature vector. We required the feature vector to contain no missing values, and thus excluded patient encounters that had one or more types of vital sign measurements missing in the observation window. The majority of the vital sign measurements, except for temperature which was sporadically measured at the bedside, were derived from high temporal resolution waveforms and matched to the temporal resolution of the corresponding measurements provided by a Garmin watch and an Oura ring. In particular, heart rate and RMSSD were calculated after extracting inter-beat interval from photoplethysmography (PPG), and respiratory rate was derived from impedance-based measurements.

The same set of demographics and vital sign features were extracted from the wearable dataset. The Oura rings provided skin temperature and RMSSD measurements. Respiratory rate was measured from the Garmin watches. Concurrent heart rate measurements from the Garmin watch and Oura ring were combined before feature extraction. Plausibility filters were applied so that unrealistic values outside of a very broad physiological range were discarded ^1^. For each subject, we extracted statistic features in 24-hour intervals within a 14-day window prior to their COVID-19 test (hence 14 observation times). Statistic features were derived from measurements collected in a 24-hour observation window prior to the observation time, similar to those used to train the hospital model. We extracted more than one day of features because we wanted to assess how early our model could detect COVID-19 infection prior to diagnostic testing. To examine the impact of daytime activity and other contextual factors on physiology, we extracted two sets of features: the first set used all vital sign measurements collected in the 24-hour observation window (“daily features”), and the second set used vital sign measurements collected during sleep in the 24-hour observation window (“sleep-only features”). Hypnogram information from the wearable devices were used to identify the sleep segments where the sleep-only features were extracted.

### Hospital model training

The model for hospital-acquired infection prediction was trained using the same methodology described previously ^15^. Specifically, we used the XGBoost algorithm ^19^ to train and test the hospital model with 5-fold cross-validation. Hyperparameters were optimized using grid search. The set of hyperparameter that yielded the best model performance averaged from the 5 validation folds were used to train the final model for assessing its performance in the wearable dataset.

### Testing the hospital model in the wearable dataset

To assess the performance of the hospital model in predicting COVID-19 infection, we defined a true positive as being a positive model prediction within the 14-day period prior to a positive COVID-19 test for the positive class, and a true negative as being a negative model prediction within the 14-day period prior to a negative COVID-19 test for the negative class. Because infection risk scores from the model were calculated in 24-hour intervals within a 14-day period, a positive model prediction was defined as one with at least one prediction within the 14-day period above the defined risk threshold, and a negative model prediction was defined as one with all predictions within the 14-day period below the defined risk threshold. In other words, we computed the hospital model outputs - which were probabilistic scores that estimated the likelihood of a given subject being infected – from the demographics and vital sign features for each day (or each sleep segment) and took the maximum score during the 14-day window for each subject. We then compared the maximum scores between COVID-19 positive and negative subjects and reported the model performance using the following metrics:

- Area under the Receiver Operating Characteristic curve (AUROC),
- Average Precision (AP),
- True Negative Rate (Specificity),
- True Positive Rate (Sensitivity, or Recall), including:

`o` Sensitivity(Break-Even): Sensitivity at the break-even point, where Sensitivity and Precision are equal,
`o` Sensitivity(80%): Sensitivity when Specificity=0.8,
`o` Sensitivity(90%): Sensitivity when Specificity=0.9.

The significance of an AUROC value was assessed by performing a permutation test. The class labels were randomly permuted 1000 times to estimate the empirical distribution of a “random” AUROC. The observed AUROC value was then compared with this bootstrapped empirical distribution to calculate the p-value.

To estimate the overall lead time of positive classification, we identified the days (interpolated) in which the hospital model prediction exceeded a predefined threshold of sensitivity = 0.6 within the 14-day window prior to COVID-19 testing. The threshold was suggested by the study principal investigators in the US DoD sites. The lead time was then defined as the average across these positive days for each user and then aggregated across the cohort for the final mean estimate of the lead time for COVID-19 classification (False Negatives have lead time of 0 days). We also overlayed risk scores with time to have a visual representation of risk score elevation during the infection period.

To reduce the impact of dataset shift on model performance, we performed a monotonic feature transformation by first calculating percentile values of each feature in the hospital dataset and the wearable dataset respectively, and then replacing wearable feature values with the hospital feature values that shared the same percentile. The percentile values of a given feature were calculated in each dataset using all samples without distinguishing between positive and negative class labels. This way, we calibrated features from wearables to match the distribution in the hospital dataset without knowledge of the class labels. We then validated the performance of the hospital model on the calibrated wearable features and compared it with model performance on wearable features before feature transformation. To understand the data requirements for feature transformation, we performed additional benchmarking experiments with restrictions on the type and size of wearable data used for feature transformation, including: using wearable data acquired when the subjects were not under impact of COVID-19 infection; using wearable data from subjects that were not used to test the model performance; using the most recent days of wearable data prior to diagnostic testing; and using wearable data from randomly down-sampled cohorts or subject days (without replacement, 10 iterations).

## RESULTS

### Cohorts and Features for Training and Testing

The cohort selection criteria for training the hospital model resulted in a total dataset size of 9,517 patient encounters with waveform recordings around the time of clinical suspicion of hospital-acquired infections (not including COVID-19). Of these patient encounters, 3,951 (3,665 controls and 286 HAIs; 51% Banner Health and 49% MIMIC-III) had overlapping PPG waveforms and impedance-based measurements with good data quality, and therefore had the full set of 18 demographics and vital sign features (see METHODS) available at 1-hour before clinical suspicion of infection. These 3,951 patient encounters were used to train the hospital model of hospital-acquired infection prediction.

The cohort selection criteria for testing the trained hospital model resulted in 301 COVID-19 positive subjects and 2,111 COVID-19 negative subjects from the wearable dataset. Within the 14-day windows prior to COVID-19 tests from these subjects, a total of 33,164 subject days and 31,269 subject sleep segments had vital sign measurements that passed our plausibility filter. From these subject days, we extracted the feature vectors comprising the same set of 18 features that was used to train the hospital model, using either all available vital sign measurements or those measured during sleep (“daily features” and “sleep-only features”, see METHODS), to quantify the performance of the trained hospital model in predicting COVID-19 infections.

### Differences between training and testing datasets

The joint distribution of inputs and outputs of the infection prediction model differed between the training scenario in the hospital dataset and the testing scenario in the wearable dataset – a problem known as “dataset shift” ^16^. Here we describe five sources of dataset shift in our study.

First, the demographics of the training and testing cohorts were different. The patients from the hospital dataset were older than the subjects from the wearable dataset (Figure 1A), and the wearable dataset had an imbalanced sex ratio than the hospital dataset (Figure 1B, 20% female in the wearable dataset versus 47% female in the hospital dataset). Both age and sex may result in differences in physiology ^20–30^.

Second, the health states of the training and testing cohorts were different. Patients in the hospital dataset are those who developed hospital-acquired infections during their stays in general wards or in some cases intensive care units, and are likely older adults with comorbidities and under medical treatments, therefore the physiological measurements in the hospital dataset were more likely to be abnormal and unstable compared to the physiological measurements in the wearable dataset where healthy young military personnel performing their daily duty were monitored. We found that patients in the hospital dataset had higher heart rate and higher respiratory rate than the subjects in the wearable dataset (see the Average and Maximum statistic feature in Supplementary Table 1), which were consistent with an overall declined health state ^30–33^. The hospital patients also had larger variations in heart rate and respiratory rate than the subjects in the wearable dataset (see the Standard Deviation statistic feature in Supplementary Table 1).

Third, the data sources where the physiological features were extracted from were different between the hospital dataset and the wearable dataset. Temperature features were extracted from core body temperatures in the hospital dataset, whereas in the wearable dataset skin temperatures measured at the fingers were used. We found that skin temperature had lower values and larger variance compared with core body temperature (Figure 1C, Supplementary Table 1), which was consistent with the literatures ^34–37^.

Fourth, the processing methods to extract physiological signals were different between the two datasets. Heart rate variability measurement RMSSD were computed based on pulse estimates of heart beats. However, the signal processing algorithms that Oura ring used could be different from ours in detecting the fiducial points on the pulse waveforms, and in the validation of the resulted inter-beat intervals. We suspected that differences in the signal processing algorithms to obtain RMSSD also contributed to the distribution differences in the RMSSD features between the hospital dataset and the wearable dataset (Supplementary Table 1), in addition to the demographics and health state differences mentioned above.

Finally, wearable physiological data is acquired in an unconstrained, real-world environment, which is influenced by everyday activities and other contextual factors. In contrast, hospital physiological data is typically acquired when the patient is sedentary. Daytime activity such as physical exercise increases heart rate and respiratory rate ^30,31^, which is a confounding factor to infection prediction because infections cause similar changes in vital signs ^32,38^. Skin temperature also changes dynamically upon physical exercise, and the directionality of change depends on the intensity level of the exercise and whether the skin temperature is measured over active or non-active muscles ^39^. When limiting feature extraction to wearable physiology data acquired during sleep, we found that sleep-only features have different data distributions compared to the daily features (Supplementary Table 1). For example, the data distribution of the mean temperature feature was shifted towards higher values when restricted to measurements during sleep (Figure 1C).

We included a full comparison of feature values in Supplementary Table 1.

**Figure 1:**
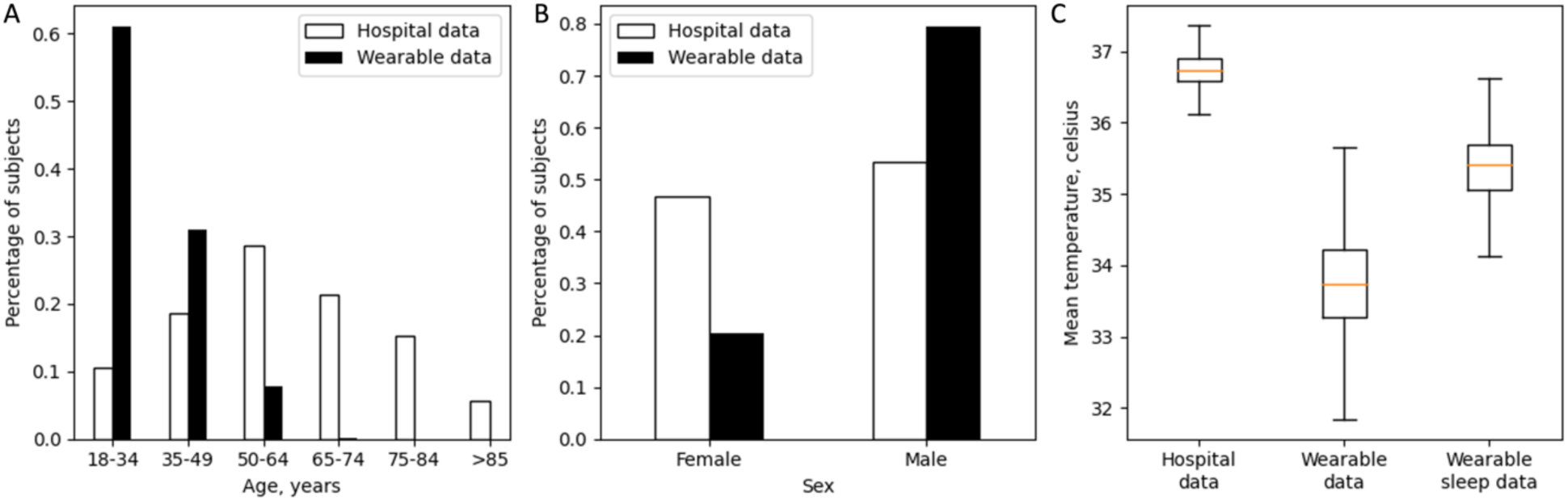
Comparison between hospital dataset and wearable dataset. (A) Age distribution of hospital dataset (white) and wearable dataset (black). (B) Sex distribution of hospital dataset (white) and wearable dataset (black). (C) Boxplot of mean temperature feature value from hospital dataset (left), wearable dataset (middle), and wearable dataset during sleep (right).

### Experiment design

We explored two approaches to correct for differences in data distributions between hospital and wearable datasets. First, we limited feature extraction to wearable physiological data from wearable sensors acquired when the subject was sleeping. This approach directly mitigated dataset shift by removing contextual confounders of daytime activities. Second, we explored a monotonic feature transformation method to convert the data distribution of physiological features in the wearable dataset to match the data distribution in the hospital dataset. This approach addressed covariate shift – one of the three types of dataset shift (see INTRODUCTION) - due to differences in demographics and health state between hospitalized patients and subjects in the wearable dataset, as well as differences in physiological measurements between COTS wearables and hospital grade devices. We compared model performances with or without using such correction techniques (Experiments I, II, III, IV in Figure 2), and in addition benchmarked data requirements (Experiments V, VI, VII):

- Experiment I: a baseline comparison where the trained hospital model was directly applied to the daily features from the wearable dataset. Physiological measurements during both awake and sleep were used to extract the daily features.
- Experiment II: the trained hospital model was tested on sleep-only features from the wearable dataset. Sleep-only features were extracted from the same window and time interval as the daily features but only using measurements during sleep segments.
- Experiment III: the trained hospital model was tested on sleep-only features after the sleep-only features were transformed to match the distribution of the hospital dataset.
- Experiment IV: the trained hospital model was tested on daily features after the daily features were transformed to match the distribution of the hospital dataset.
- Experiments V, VI, VII: benchmarking the amount and type of wearable data needed for the monotonic feature transformation.

**Figure 2:**
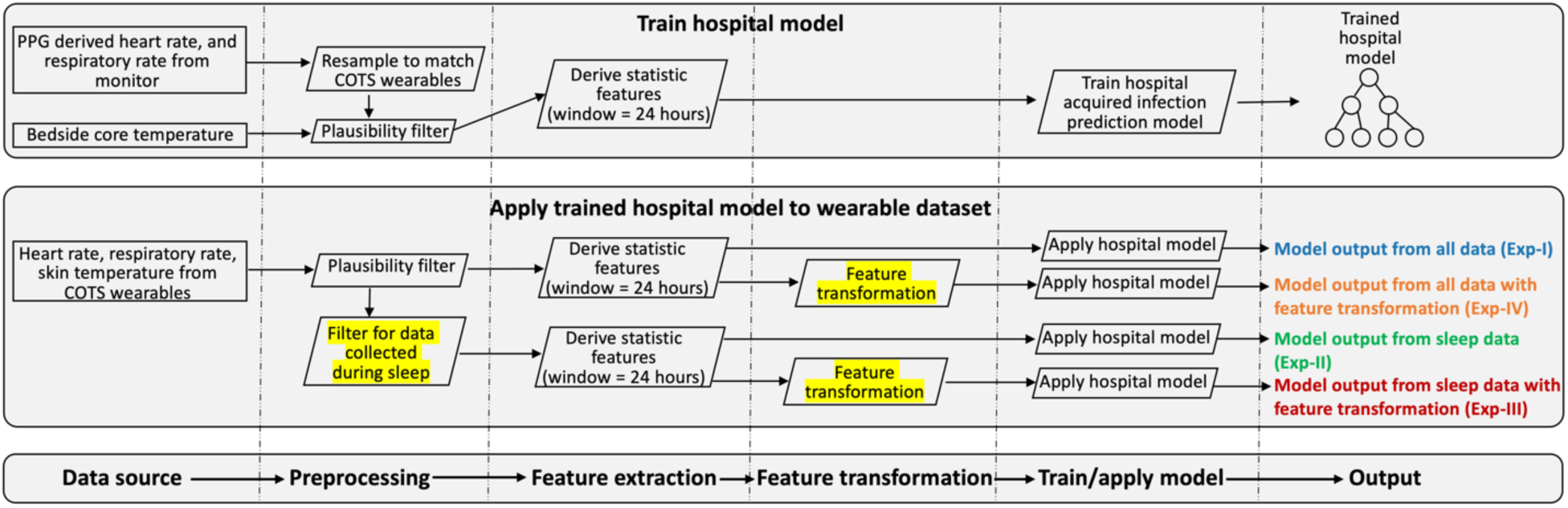
Schematic view of pipelines for training the hospital model (top box) and for testing the trained model in the wearable dataset (middle box). Similar steps of the two pipelines are aligned (bottom box). The trained hospital model was applied to the wearable dataset with or without the two dataset shift corrections (highlighted, middle box), which resulted in four experiments (Exp-I, II, III, IV in middle box) to compare model performance.

### Baseline comparison (Experiment I)

We directly applied the hospital model trained for hospital-acquired infection prediction to the wearable daily features and quantified its performance in predicting COVID-19 infections. We hypothesized that the hospital model would not generalize well in predicting COVID-19 infections, due to the differences between hospital and wearable physiological feature spaces. We found that the hospital model performed at Area under ROC Curve (AUROC) = 0.527, Average Precision (AP) = 0.132, Sensitivity = 0.163 and Specificity = 0.866 at break-even point, Sensitivity = 0.193 and 0.113 respectively when Specificity was at 0.8 and 0.9. This performance was at chance level (p=0.07 for AUROC), suggesting that the hospital model failed to generalize when directly applied to wearable dataset.

### Removing contextual confounders (Experiment II)

When controlling for contextual factors like daytime activity, we found that the hospital model using the sleep-only features performed at AUROC = 0.644 (p<0.001), AP = 0.260, Sensitivity = 0.279 and Specificity = 0.897 at break-even point, Sensitivity = 0.402 and 0.269 respectively when Specificity was at 0.8 and 0.9. Thus, using sleep-only features resulted a 22% boosting of performance in terms of AUROC, suggesting the importance of controlling for contextual confounders when extracting the likelihood of infection from wearables physiological data.

### Applying feature transformation after removing contextual confounders (Experiment III)

We hypothesized that a monotonic feature transformation procedure which transforms the wearable feature values to match the distribution in hospital dataset (see METHODS) could improve performance of the hospital model. Using mean temperature feature as an example (Figure 3), the feature transformation procedure based on matching feature values that share the same 0-100 percentile value in their corresponding datasets resulted in an almost identical data distribution of the mean temperature feature between the two datasets, despite large discrepancies in the data distributions before transformation. Hence, we performed the same feature transformation procedure independently on each feature, and evaluated the performance of hospital model on the wearable dataset after all the features were transformed. We found that the hospital model performed at AUROC = 0.740 (p<0.001; Figure 4A, red), AP = 0.330 (Figure 4B, red), Sensitivity = 0.379 and Specificity = 0.910 at break-even point, Sensitivity = 0.588 and 0.409 respectively when Specificity was at 0.8 and 0.9, using transformed wearable sleep-only features. Applying feature transformation on the sleep-only features resulted an additional 15% boosting of performance in terms of AUROC (0.740 versus 0.643, red versus green in Figure 4A).

**Figure 3:**
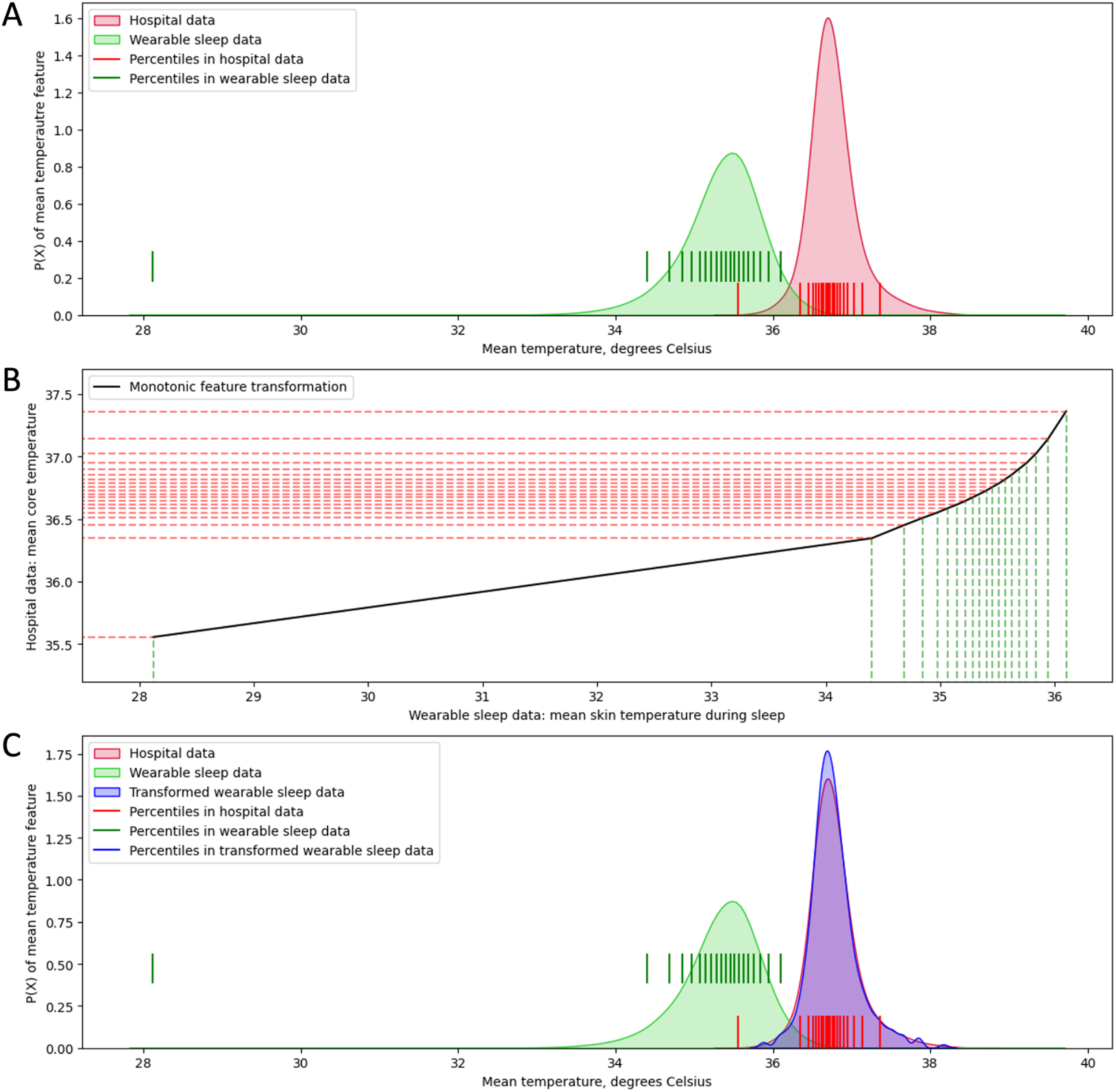
Monotonic feature transformation of mean temperature feature. Red, hospital dataset; green, wearable dataset (sleep-only features); blue, transformed wearable dataset (sleep-only features). (A) Data distribution of mean temperature feature: red and green shaded areas describe data distribution from hospital and wearable sleep data respectively. Vertical lines mark the 0-100 percentile values in 5% intervals on the x-axis corresponding to each dataset. (B) Monotonic feature transformation curve (black) where feature values with the same percentile value are mapped between two datasets. Dashed lines mark the 0-100 percentile values in 5% intervals on the x-axis for wearable sleep data (green) and on the y-axis for hospital data (red). (C) Data distribution of mean temperature feature: red, green and blue shaded areas describe data distribution from hospital dataset, wearable sleep dataset and transformed wearable sleep dataset respectively. Vertical lines mark the 0-100 percentile values in 5% intervals on the x-axis corresponding to each dataset; blue vertical lines are overlapped with red vertical lines.

### Applying feature transformation without removing contextual confounders (Experiment IV)

We further investigated whether the same feature transformation procedure could improve the performance of the hospital model on wearable features without removing the contextual confounder of awake versus sleep. Similarly, we calculated percentile values of daily wearable features derived from awake and sleep data combined, replaced the feature value with the corresponding value from the hospital dataset, and evaluated the performance of the hospital model on the transformed features. The model had an AUROC = 0.566 (p<0.001; Figure 4A, orange), AP = 0.158 (Figure 4B, orange), Sensitivity = 0.256 and Specificity = 0.844 at break-even point, Sensitivity = 0.296 and 0.146 respectively when Specificity was at 0.8 and 0.9, when applied to the transformed wearable features without using sleep data exclusively. The model performance was slightly better than before feature transformation (AUROC: 0.565 versus 0.526, orange versus blue in Figure 4A), but the improvement was not as substantial as when applying feature transformation to the sleep-only features (AUROC: 0.740 versus 0.643, red versus green in Figure 4A). These results suggested that both controlling for contextual cofounders and applying feature transformation to address dataset shift were important to enable good model performance.

### Comparison with previous work

We have shown that the hospital model trained for hospital-acquired infection prediction performed the best in detecting early signs of COVID-19 infection on wearable dataset when feature transformations were performed and when only sleep data were considered (AUROC = 0.740, Experiment III). Although this performance is viable for a system, it was lower than our previously reported solution using a model trained directly on wearable dataset with COVID-19 labels (AUROC = 0.82) ^1^. This was expected because the hospital model was designed to be an economical minimal viable solution that uses no COVID-19 labels for training, and thus not capable of controlling for concept drifts and/or label shifts. When overlaying risk scores with time from the Experiment III hospital model, on average subjects with positive COVID-19 test results showed risk score elevations around COVID-19 test time (Figure 4C, black), whereas subjects with negative COVID-19 test maintained their baseline risk scores (Figure 4C, blue). Based on a cut-off risk threshold of 15 (yielding 60% sensitivity and 78% specificity), we identified the days in which the model output exceeded the defined threshold within the 14-day window prior to COVID-19 testing to estimate the lead time of positive classification (see METHODS). We found that the Experiment III hospital model successfully predicted COVID-19 infection, on average, 2.2 days prior to testing. This lead time was slightly lower but comparable to our previously reported wearable solution of 2.3 days prior to testing ^1^.

**Figure 4:**
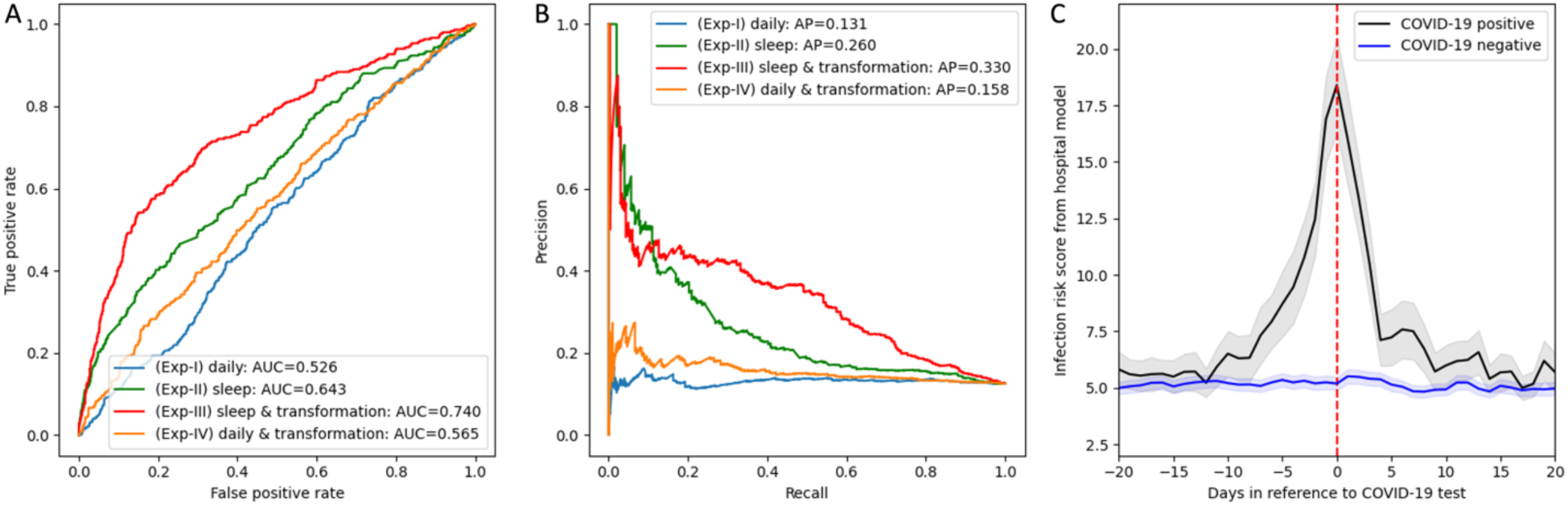
Hospital model performance and risk scores in detecting COVID-19 infection from wearable dataset. (A) Receiver Operating Characteristic (ROC) curves. Experiment I (blue): hospital model directly applied to wearable daily features. Experiment II (green): hospital model applied to wearable sleep-only features. Experiment III (red): hospital model applied to wearable sleep-only features after feature transformation. Experiment IV (orange): hospital model applied to wearable daily features after feature transformation, without using sleep data exclusively. Area under the ROC curve (AUROC) for each experiment is included in the figure legend. (B) Precision-recall curve. Colors are the same as described in subplot A. Average Precision (AP) score for each experiment is included in the figure legend. (C) Mean infection risk score based on the output of the best generalized hospital model (Experiment III: sleep-only features + feature transformation) in 301 COVID-19 positive subjects (black) and 2,111 COVID-19 negative subjects (blue) as a function of number of days relative to the COVID-19 test time (red). Grey and light-blue shaded area depicts 95% confidence interval.

### Data requirements for feature transformation (Experiments V, VI, VII)

Given that our best generalized hospital model (Experiment III) performed reasonable but inferior to our previous wearable model ^1^, it is most sensible to use a hospital model for predicting COVID-19 in the absence of the wearable model, e.g. at the onset and during the early stage of the outbreak when data from COVID-19 positive cases were limited or unavailable to train a wearable model. Therefore, we investigated the data requirements of the generalized hospital model in Experiment III, in particular, the type and amount of wearable sleep data needed for the feature transformation. A favorable solution should require minimal COVID-19 positive instances. We performed three sets of additional experiments.

First, we asked whether illness data of COVID-19 were required for feature transformation (Experiment V). Interestingly, we found that baseline healthy data was sufficient because 1) using wearable sleep data from subjects that only reported negative test results for the feature transformation resulted in similar AUROC of 0.741 (Experient V-a, Supplementary Table 2), and 2) using wearable sleep data 4 weeks to 2 weeks before COVID-19 test – a time range when subjects were not infected - achieved similar results (AUROC = 0.741; Experient V-b, Supplementary Table 2).

Second, we asked whether wearable sleep data for feature transformation needed to be from the same subjects (Experiment VI, Supplementary Table 2). We randomly split the subjects in the wearable dataset into 5 folds, and for each subject we used subjects from the other four folds to transform the features of the given subject. The model performed with an AUROC of 0.741, suggesting that the wearable data used for feature transformation do not need to come from the same subjects used to test the model.

Third, we examined the minimum sleep data needed for feature transformation by benchmarking model performance against using sleep data from reduced number of days or from reduced number of subjects (Experiment VII). We gradually decreased the number of days from the 14 days prior to COVID-19 test where the wearable data were used for feature transformation (Experiment VII-a, Supplementary Table 2). We found that the model performed at AUROC of 0.74 when more than 2 days immediately preceding the COVID-19 test were used for feature transformation, and the model performed at AUROC = 0.73 when using data from the day before or two days before COVID-19 test for feature transformation. We also benchmarked against data from randomly selected days within the 14-day window prior to the COVID-19 test for feature transformation and found that the model performed at AUROC of 0.74 for all experiments - randomly selecting number of N days where N ranges from 1 to 13 days (Experiment VII-b, Supplementary Table 2). Further, we pooled all subject days and used random down-samples for feature transformation (Experiment VII-c, Supplementary Table 2). We found that the model performed at AUROC of 0.74 for all experiments of reduced subject days (number of reduced subject days: 25,000, or 20,000, or 15,000, or 10,000, or 7,500, or 5,000, or 3,000, or 1,000, or 500, or 300), even when only 300 subject days were used. Regarding the number of subjects needed, we used data from randomly down-sampled subjects for feature transformation and found that the model performed at AUROC of 0.74 for all experiments of reduced number of subjects (number of reduced subjects: 2,000, or 1,500, or 1,000, or 500, or 250, or 100, or 50, or 25), even when the number of subjects was reduced to 25 (Experiment VII-d, Supplementary Table 2).

Summarizing all the experiments (Supplementary Table 2), we concluded that healthy wearable data from 25 subjects collected in a period of 14 days for feature transformation would be sufficient to ensure the same model performance of AUROC = 0.74.

## DISCUSSIONS

This study demonstrated the feasibility of applying a machine learning model trained on hospital data to detect early signs of COVID-19 infection in physiological data from COTS wearables outside of hospitals. Our hospital model was trained from hospitalized patients and vital signs collected from hospital grade devices to test against a set of common hospital-acquired infections (prior to the COVID-19 outbreak), therefore had no prior knowledge of COVID-19 infections and no exposure to physiological data collected through COTS wearables. Nevertheless, after controlling for dataset shift, the hospital model performed at AUROC = 0.74 in alerting COVID-19 infection before diagnostic testing from wearable physiology monitoring in military personnel under unrestrained use. This performance was lower than our previously reported solution using a model trained directly on wearable dataset with COVID-19 labels ^1^, but is nevertheless viable for a system, and can detect COVID-19 infection 2 days before diagnostic testing, with no need of model retraining. Importantly, our approaches in addressing dataset shift did not require any labeled data of COVID-19 cases; rather, a small dataset from healthy subjects – e.g. 2 weeks of wearable data from 25 subjects – was sufficient to generalize the hospital model to predict COVID-19 infection from wearable data with an AUROC of 0.74. Therefore, our efficient solution of generalizing the hospital model of infection prediction to wearable physiological monitoring would be most economical and useful at early onset of outbreaks of novel infections when data from positive cases are limited or unavailable to train an pathogen-specific model – such as our previously reported COVID-19 wearable model ^1^. Because a small amount of healthy baseline data is feasible to collect prior to any infection outbreak, the transformation function to calibrate the feature values can be derived to enable rapid deployment of a pre-trained model. We anticipate such a solution could create a big impact in infectious disease control, as transmission prevention at the onset and during the early outbreak of an infectious disease is critical.

The two enablers of our solution of generalized hospital model were 1) the isolation of contextual confounders, focusing on sleep-only wearable data, and 2) feature transformations that calibrated the wearable feature values to match the distribution of the hospital model training data and that do not rely on positive labels. Both reduced the differences in the joint distribution of the physiological features X and the infectious disease labels Y between the hospital dataset and the wearable dataset, therefore mitigating dataset shift. The model performed at chance level without these two corrections and performed at AUROC of 0.74 when and only when both corrections were used. This is likely because the two methods controlled for different aspects of dataset shift. Feature transformation is a correction technique for covariate shift (see INTRODUCTION) because it modifies the probability distribution of the physiological features P(X). Removing contextual confounder of daytime activities, on the other hand, controls for both covariate shift P(X) and to some extent concept drift P(Y|X). For example, increases in heart rate in hospitalized patients are associated with increased risk of infection ^38^; in contrast, increases of heart rate in the subjects from the wearable monitored cohort could be normal physiology change, e.g. if the subjects are exercising ^30^. Therefore, daytime activities such as physical exercises affect the wearable physiology data in such a way that increases the likelihood that they will be misclassified by the hospital model as infection cases. Hence it is beneficial to use sleep-only features in our study, and that it is not sufficient to perform feature transformations on the daily features without isolating sleep periods.

In our study we used hypnogram information from wearables to identify measurements during sleep to compute sleep-only features so that both P(X) and P(Y|X) were more similar to the hospital dataset, where the physiological measurements were acquired when patients were sedentary. We could also apply the hospital model to the wearable dataset in other similar scenarios such as during wakefulness, but limited to resting/sedentary states. It is possible that there are other contextual confounders that we could identify and isolate from the wearable dataset to further improve the model performance of the generalized hospital model. Identifying contextual factors does not require any explicit knowledge of data distributions of the training nor testing datasets but replies on domain knowledge of the model training and application scenarios. Removing contextual factors, however, relies on the availability of data elements that can be used to isolate the contextual factors.

It is challenging to address all aspects of dataset shift. In particular, label shift and concept drift would require labels to be properly addressed. Previous work that corrected dataset shift using unlabeled data typically addressed covariate shift, and involved re-training using resampling weights that were either estimated from the biasing densities ^40–42^ or inferred by comparing nonparametric distributions between training and testing samples ^43^. In contrast, the monotonic feature transformation technique described in our study requires no labels, no re-training, and is a straightforward mathematical operation that preserves the rank order of data but modifies the shape of the distribution. By doing so, we are minimizing the dataset differences in physiological signals caused by the differences in individual and group baselines, and by the differences in measurement devices, yet preserving the relative rank of infection risk among individuals. Our hospital model was based on ensembles of decision tree which makes aggregated decisions from individual features on each tree split. This makes it possible for us to manipulate the distribution of each feature independently without altering the overall decision from the tree ensembles based on the feature ranks (e.g. P(Y|X) is unchanged for monotonic transformations of X, where X is the physiological features and Y is the infection labels). Algorithms based on decision trees are particularly suitable for disease modeling, as typically lower and/or higher clinical measurements are associated with declined health. In other words, infection risk as a function of clinical measurements resembles a U-shape curve or a monotonic function. This is the reason why preserving the rank of feature values worked in our solution as it preserved the rank of infection risk, e.g. both a high rank of skin temperature and a high rank of core temperature are associated with high infection risk, therefore the conditional probability of COVID-19 infection risk given skin temperature P(Y_covid|X_skin) can be monotonically mapped to the conditional probability of infection risk given core temperature P(Y_infection|X_core).

Our feature transformation technique requires no labels (therefore is “unsupervised”), no re-training, and is computationally inexpensive and interpretable, compared with previous work that corrected dataset shift ^40,41,41–43^. It is device-agnostic by nature, and we demonstrated its effectiveness in addressing the dataset shift due to differences in measurement devices, e.g. the skin temperature feature from Oura ring was transformed to have almost identical distribution as the core temperature feature from hospital grade device (Figure 3). Removing context confounders have its challenges in first identifying the relevant context and then finding data elements that can be used to isolate the context, but theoretically has the potential to make our solution context-agnostic. Our generalized hospital model of infection prediction performed well in detecting COVID-19, despite pathogen differences in COVID-19 infection and the set of hospital-acquired infections used to train the hospital model. Therefore, we believe the generalized hospital model can be easily adapted to deploy in other scenarios of infection prediction, and it is not restricted to a specific set of wearable devices, a specific population, or a specific context. For example, the hospital model of infection prediction may be used to track the health state of healthcare professionals during flu season with a different set of wearables, given that similar types of vital sign signals are collected, and appropriate dataset shift transformations are applied.

## CONCLUSTIONS

We found that an infection prediction model developed for hospitalized patients can detect early signs of COVID-19 infection from wearable physiological monitoring (AUROC=0.74), on average 2 days earlier than diagnostic testing, provided that a small sample (e.g. 25 subjects in a period of 14 days) of wearable data from healthy subjects is available to address the dataset shift between hospital dataset and wearable dataset, and that sleep markers can be extracted to control for contextual effects in wearable dataset. Our approaches to transform features between datasets and isolate contextual confounders can enable rapid deployment of a pre-trained infection prediction model at the onset of novel infection outbreaks.

## Data Availability

MIMIC-III dataset is available in PhysioNet repository, https://mimic.physionet.org/. The Banner Health dataset is a proprietary dataset that is not publicly shareable. The wearable dataset is from US military personnel and is not publicly shareable.

## ACKNOWLEDGEMENTS

This study is sponsored by the US Department of Defense (DoD), Defense Threat Reduction Agency (DTRA) under contracts: W15QKN-18-9-1002 (CB10560), HDTRA1-20-C-0041, HDTRA121C0006. The views, opinions and/or findings expressed are those of the authors and should not be interpreted as representing the official views or policies of the Department of Defense or the US Government. We appreciate the vision, leadership, and sponsorship from the US Department of Defense and the US Government: Edward Argenta, Christopher Kiley and Katherine Delaveris. We recognize our former Philips North America colleague Saeed Babaeizadeh for PPG signal processing.

## LIST OF ABBREVIATIONS

AI: Artificial Intelligence
ML: Machine Learning
CDS: Clinical Decision Support
Spec: Specificity
Sens: Sensitivity
AUROC: Area under the Receiver Operating Characteristic Curve
AP: Average Precision
COTS wearables: Commercial-off-the-shelf wearables
HAI: Hospital-acquired Infection

## DECLARATIONS

### Ethics approval and consent to participate

The MIMIC-III project was approved by the Institutional Review Boards of Beth Israel Deaconess Medical Center and the Massachusetts Institute of Technology. Banner Health data use was a part of a retrospective deterioration detection study approved by the Institutional Review Board of Banner Health and by the Philips Internal Committee for Biomedical Experiments. For both hospital datasets, requirement for individual patient consent was waived because the project did not impact clinical care, was no greater than minimal risk, and all protected health information was removed from the limited dataset used in this study.

### Conflicts of Interest Statement

Authors TF, SM, BC, RD and IS are employees of Philips North America. Author DM was employee of Philips North America. Author DS is employee of Banner Health. All authors declare no other competing interests.

### Funding Statement

This study is sponsored by the US Department of Defense (DoD), Defense Threat Reduction Agency (DTRA) under contracts: W15QKN-18-9-1002 (CB10560), HDTRA1-20-C-0041, HDTRA121C0006. The funding body did not play a role in the study design, collection, analysis, interpretation of data, the writing of this article or the decision to submit it for publication. The views, opinions and/or findings expressed are those of the authors and should not be interpreted as representing the official views or policies of the Department of Defense or the US Government.

### Authors’ contributions

TF, DM and BC participated in the conception of the study. TF analyzed the data, trained, validated the models, and wrote the first draft. SM extracted waveform numeric, processed PPG waveforms and extracted heart rate variability features. RD extracted labels from the wearable dataset. BC and IS set up the ETL pipeline for wearable data processing. DS provided clinical consultation and reviewed the manuscript. All authors participated in interpretating the results, writing, and revising the manuscript. All authors have read and approved the manuscript.

---

**Supplementary Table 1:**
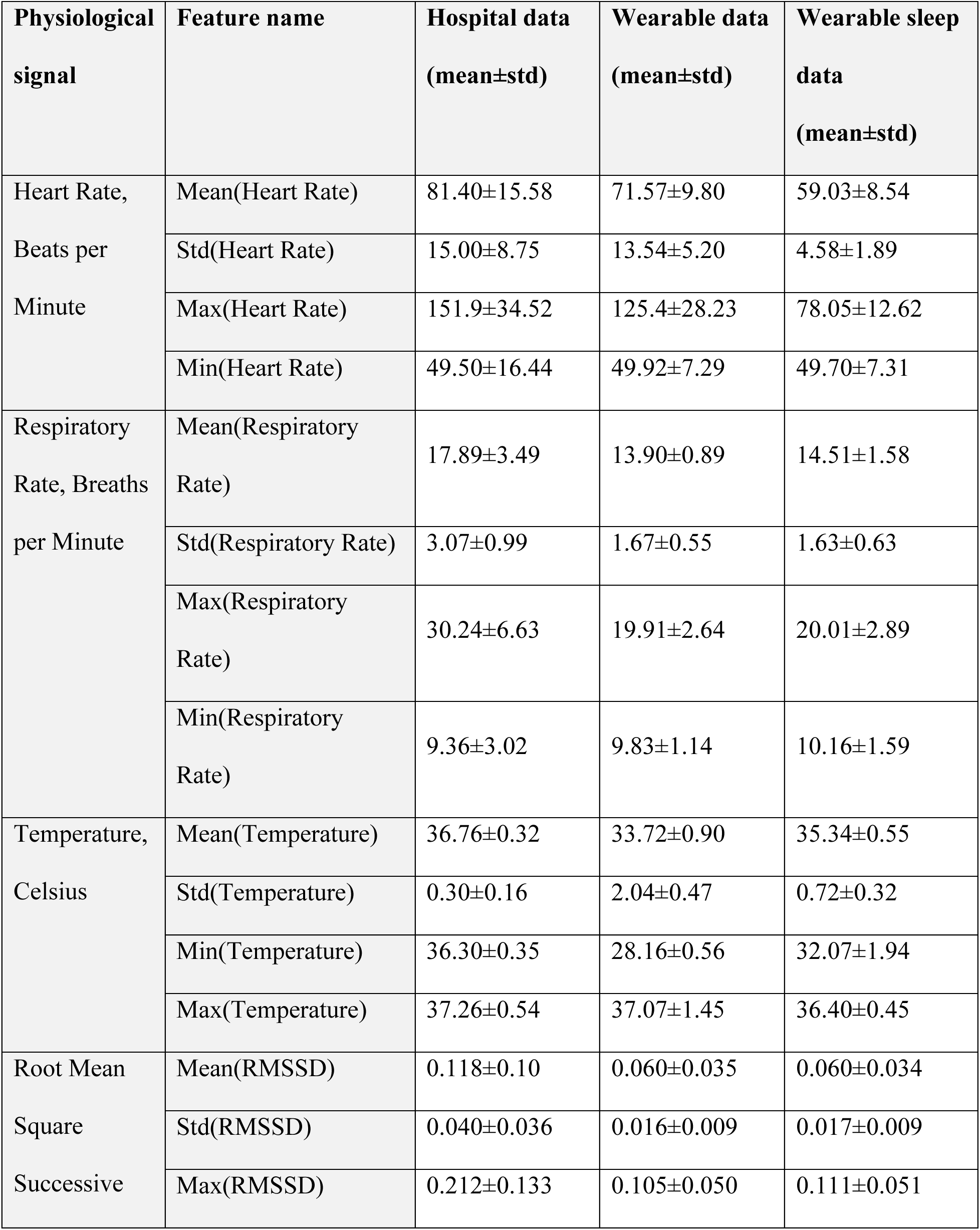
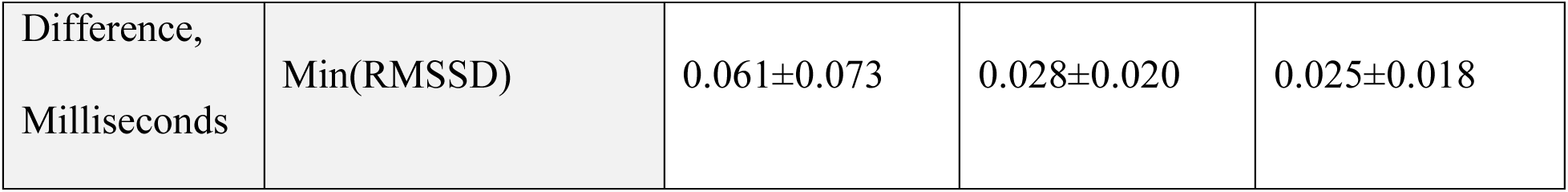
Mean and standard deviation (std) of the feature values by dataset. Hospital data – from 9,517 hospitalized patients. Wearable data – from 33,164 subject days. Wearable sleep data – from 31,269 subject sleep segments.

**Supplementary Table 2:**
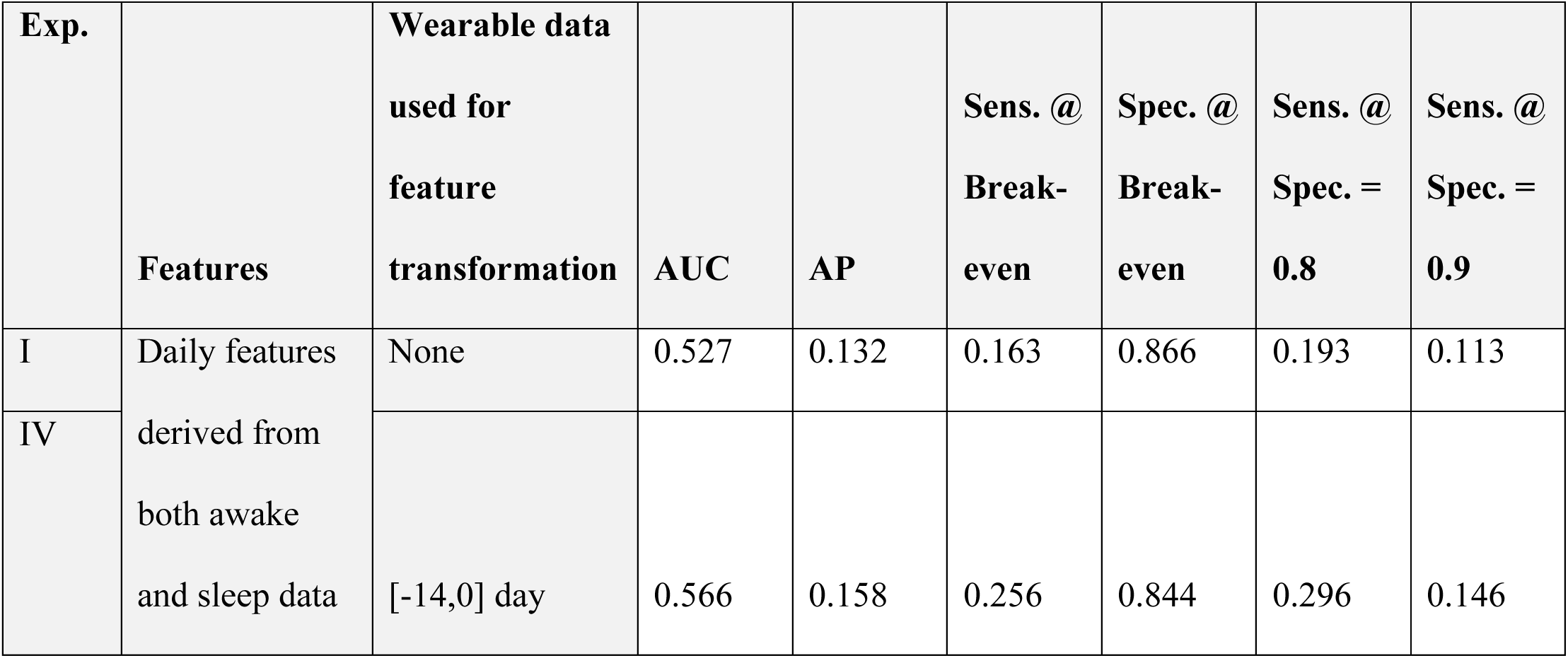
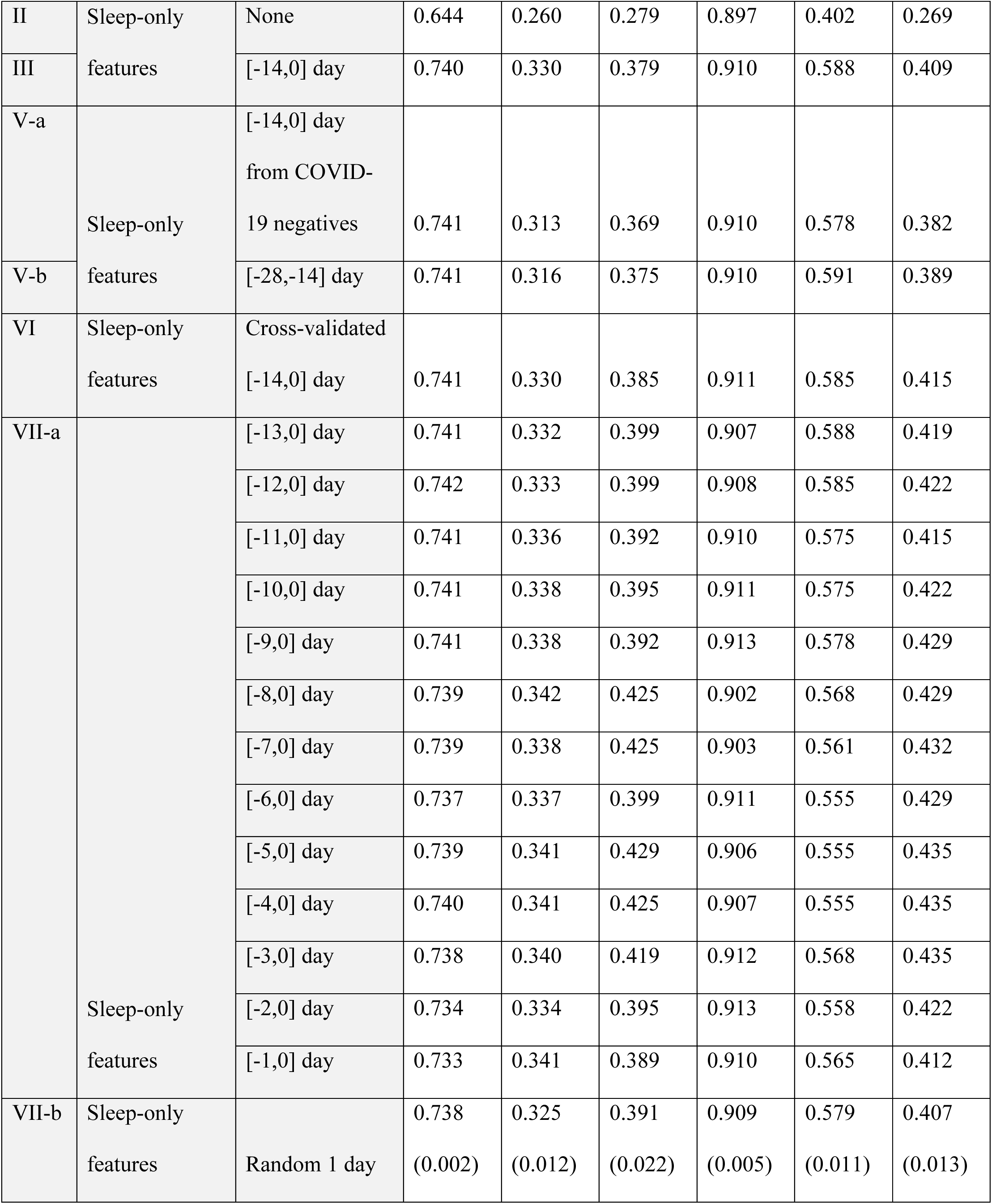
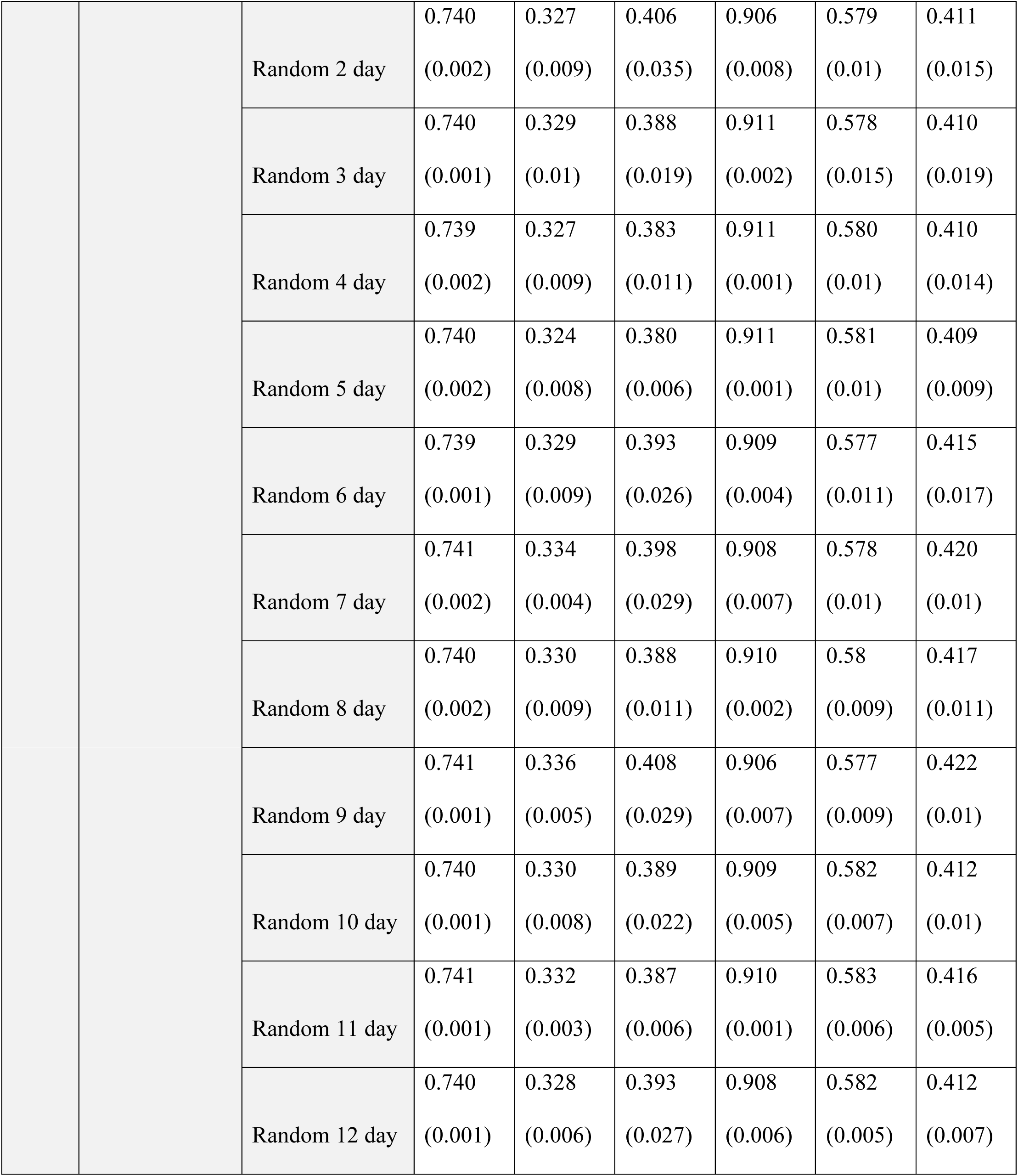
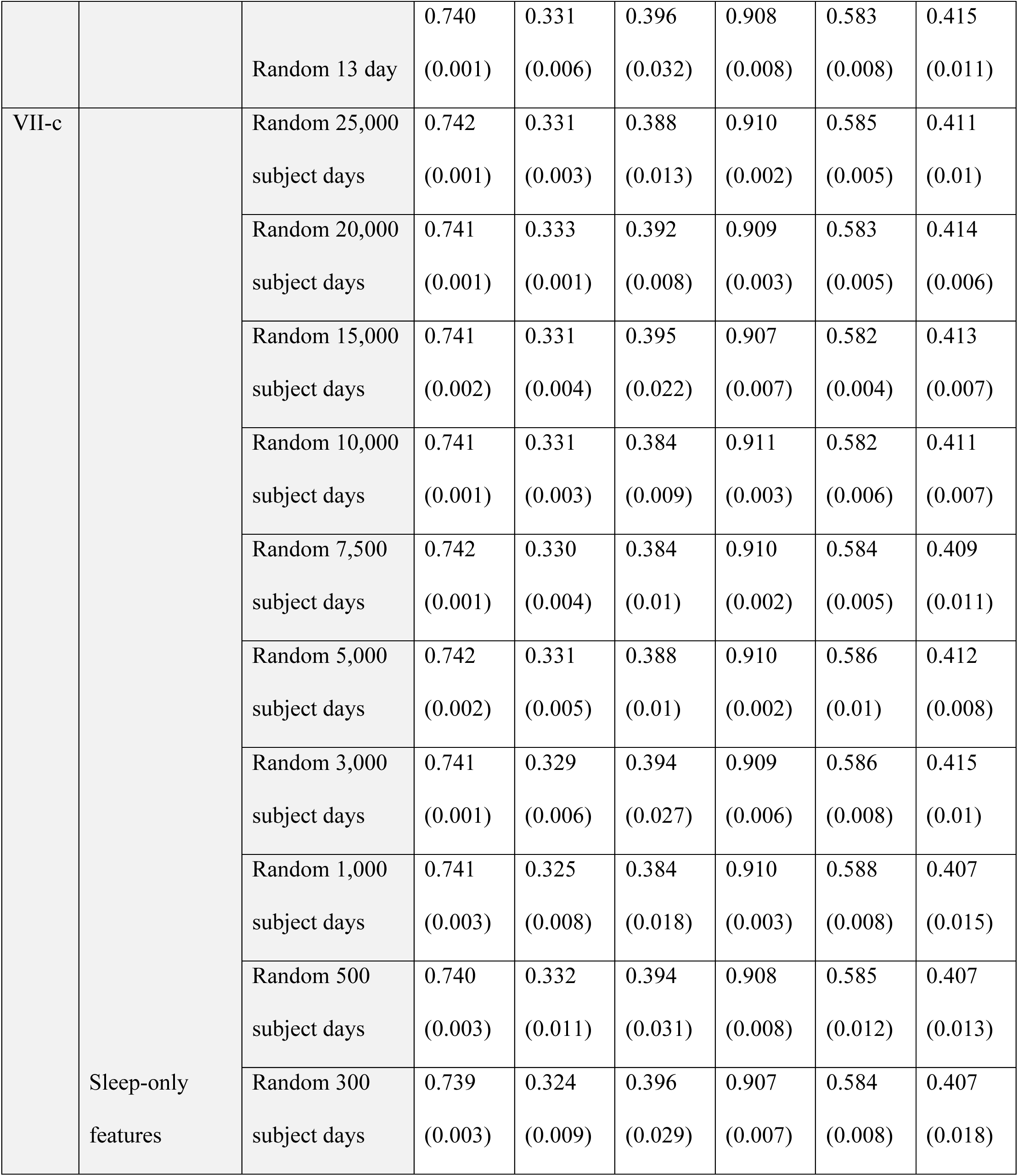
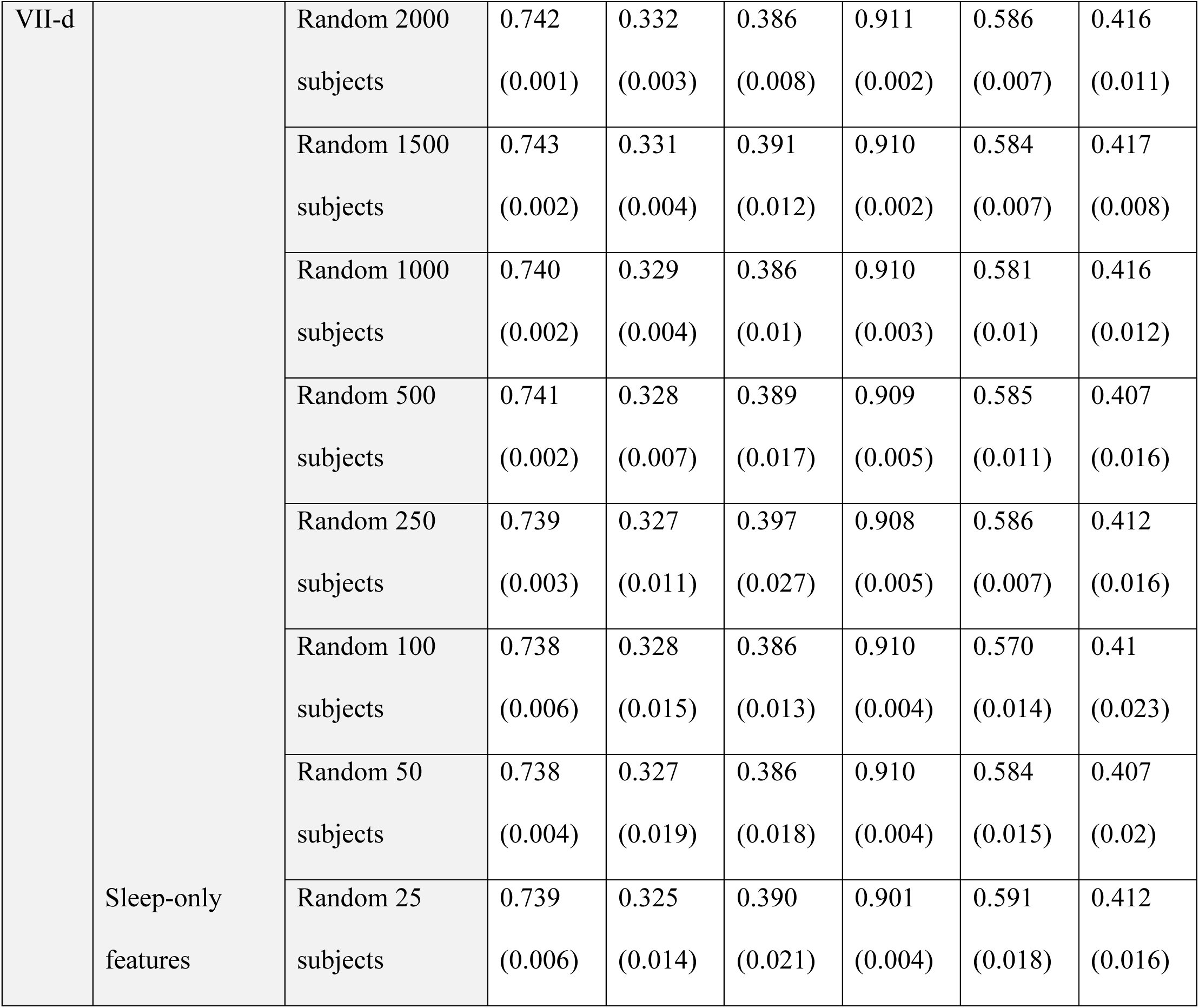
model performance. Six performance metrics were calculated: AUC (Area under ROC Curve), AP (Average Precision), Sens.@Break-even (Sensitivity at Precision-Recall break-even point), Spec.@Break-even, (Specificity at Precision-Recall break-even point), Sens.@Spec.=0.8 (Sensitivity when Specificity is 0.8), Sens@Spec.=0.9 (Sensitivity when Specificity is 0.9). Experiment I, hospital model directly applied to wearable daily features. Experiment IV, hospital model applied to wearable daily feature after feature transformation. Experiment III, hospital model applied to wearable sleep-only feature. Experiment IV, hospital model applied to wearable sleep-only feature after feature transformation. Experiment V-a, hospital model applied to wearable sleep-only feature with feature transformation using data from only subjects who reported negative test. Experiment V-b, hospital model applied to wearable sleep-only feature with feature transformation using data collected 28 days before to 14 days before COVID-19 test. Experiment VI, hospital model applied to wearable sleep-only feature with cross-validated feature transformation. Experiment VII-a, hospital model applied to wearable sleep-only feature with feature transformation using data from the recent n days prior to testing (n ranges from 1 to 13). Experiment VII-b, hospital model applied to wearable sleep-only feature with feature transformation using data from randomly selected number of n days within 14 days prior to testing (n ranges from 1 to 13), mean(std) from 10 iterations is shown. Experiment VII-d, hospital model applied to wearable sleep-only feature with feature transformation using data from randomly selected number of n subject days (n=[25000,20000,15000,10000,7500,5000,3000,1000,500,300]), mean(std) from 10 iterations is shown. Experiment VII-d, hospital model applied to wearable sleep-only feature with feature transformation using data from randomly selected number of n subjects (n=[2000,1500,1000,500,250,100,50,25]), mean(std) from 10 iterations is shown.

## Notes

### Author Declarations

The MIMIC-III project was approved by the Institutional Review Boards of Beth Israel Deaconess Medical Center and the Massachusetts Institute of Technology. Banner Health data use was a part of a retrospective deterioration detection study approved by the Institutional Review Board of Banner Health and by the Philips Internal Committee for Biomedical Experiments. For both hospital datasets, requirement for individual patient consent was waived because the project did not impact clinical care, was no greater than minimal risk, and all protected health information was removed from the limited dataset used in this study. The collection and use of the wearable dataset was approved by the Institutional Review Boards of the US Department of Defense. Informed consent was obtained from all participants.

